# Behavioral Telemetry in the ICU: Missing Orientation Assessment Predicts Mortality in Patients with Low Acute Physiologic Derangement

**DOI:** 10.64898/2026.02.23.26346916

**Authors:** Greg Born

## Abstract

**Background:** Behavioral telemetry—the analysis of clinical actions NOT taken—may identify care process failures associated with adverse outcomes. While missed nursing care predicts outcomes in survey-based studies, objective EHR-derived measures are lacking. We hypothesized that missing routine cognitive assessment in ICU patients with low acute physiologic derangement would predict mortality independent of illness severity.

**Methods:** Retrospective cohort study using MIMIC-IV (2008-2022, Beth Israel Deaconess Medical Center) with external assessment of documentation practices in eICU (208 US hospitals). We identified ICU admissions with SOFA 0-2 (low acute physiologic derangement), excluding neurological ICUs. Orientation documentation was classified within 24 hours. Primary outcome was in-hospital mortality. Multivariable logistic regression adjusted for age, sex, SOFA, and Charlson Index.

**Results:** Among 46,004 ICU patients with SOFA 0-2, 4,737 (10.3%) had no orientation documentation within 24 hours. These patients had 24.68% mortality versus 7.57% early-assessed and 4.56% late-assessed. After adjustment, missing orientation was associated with 4.29-fold higher odds of death (95% CI 3.95-4.65; E-value 8.0). In SOFA=0 patients (N=23,670), the signal strengthened (OR 5.65, 95% CI 5.03-6.35; E-value 10.8). Late-assessed patients had the LOWEST mortality (OR 0.65), arguing against reverse causation. Patients without orientation had 22% MORE chart events (1,600 vs 1,309), arguing against neglect. External assessment revealed that among 166 eICU hospitals with ≥100 eligible patients, only 5% documented orientation routinely—92% lack the infrastructure to detect this signal.

**Conclusions:** In ICU patients with low acute physiologic derangement, absence of orientation assessment is associated with 4-6 fold increased mortality. This association may identify care process failures not captured by severity scores, though prospective studies are needed to establish causality.

**What is Already Known on This Topic:** Missed nursing care—care omissions—predicts patient mortality in survey-based studies. Nurse staffing ratios are associated with mortality, but the mechanism is poorly understood. No objective, EHR-derived measure exists to detect care process omissions in real time.

**What This Study Adds:** First EHR-based operationalization of the missed nursing care construct, enabling objective, real-time detection. Missing orientation assessment associated with 4-6 fold increased mortality (OR 4.29 in SOFA 0-2; OR 5.65 in SOFA=0). Signal strengthens in SOFA=0 patients (E-value 10.8), suggesting finding is not driven by acute illness severity. Argues against reverse causation: late assessment has BETTER outcomes than early or no assessment. Argues against neglect: patients without assessment had MORE documentation, not less. Argues against immortal time bias: Never Documented patients had LONGER ICU stays (7.58 vs 3.09 days). Quantifies association: 10.3% of patients account for 27.2% of deaths. Reveals systemic gap: 92% of US ICUs lack the documentation infrastructure to detect this signal.

**Key Points:** *Question:* Does absence of routine orientation assessment predict mortality in ICU patients with low acute physiologic derangement (SOFA 0-2), independent of illness severity?

*Findings:* In this cohort study of 46,004 ICU patients with SOFA 0-2, those without orientation documentation within 24 hours had 4.29-fold higher adjusted odds of death (95% CI 3.95-4.65). In SOFA=0 patients, the signal strengthened to OR 5.65 (E-value 10.8). Patients assessed late (6-24h) had the LOWEST mortality (OR 0.65), arguing against reverse causation. Among 166 eICU hospitals, only 5% document orientation routinely— 92% lack the infrastructure to detect this signal.

*Meaning:* Missing routine cognitive assessment may identify care process failures associated with increased mortality. The finding that 92% of US ICUs lack the documentation infrastructure to detect this signal reveals a systemic gap in care process monitoring.

## Introduction

Traditional early warning systems rely on physiological parameters—vital signs, laboratory values, severity scores. These measure what clinicians observe. We propose that what clinicians fail to observe may also be predictive of adverse outcomes.

The concept of missed nursing care—defined as any aspect of required patient care that is omitted or delayed—has been established for over two decades. Kalisch and colleagues developed the MISSCARE framework demonstrating that task omissions predict patient outcomes including falls, medication errors, and mortality.^1,2^ Ball and colleagues showed that missed care mediates the relationship between nurse staffing and patient outcomes across nine countries.^3^ Aiken’s landmark work established that each additional patient per nurse increases mortality by 7%.^4,5^ However, existing research relies primarily on nurse self-report surveys, limiting real-time detection and objective measurement.

We hypothesized that electronic health record data could serve as an objective proxy for care omissions. Orientation assessment—asking patients if they know their name, location, and date—is a routine nursing assessment in the ICU that requires bedside presence, verbal engagement, and cognitive evaluation. Its absence in patients with low acute physiologic derangement may indicate a care environment where task completion has replaced cognitive engagement—and where early signs of deterioration go undetected.

We introduce the term “behavioral telemetry” to describe this approach: the systematic detection of clinical actions NOT taken, analogous to how physiological telemetry monitors biological signals. Just as an absent heartbeat triggers alarm, an absent expected assessment may signal care process failure.

We tested whether missing orientation assessment predicts mortality in ICU patients with low acute physiologic derangement, hypothesizing that: (1) absence of assessment would be associated with increased mortality after adjustment for illness severity; (2) late assessment would NOT show increased mortality, ruling out reverse causation; and (3) patients without assessment would not have reduced documentation volume, ruling out neglect.

## Methods

### Study Design and Reporting

This retrospective cohort study used MIMIC-IV v3.1 (Medical Information Mart for Intensive Care), comprising ICU admissions to Beth Israel Deaconess Medical Center from 2008-2022.^6^ External assessment of documentation practices used the eICU Collaborative Research Database (200,859 ICU stays across 208 US hospitals, 2014-2015).^7^ The study followed STROBE guidelines.^8^ MIMIC-IV and eICU are publicly available de-identified databases; institutional review board approval was obtained by the database maintainers. Individual patient consent was waived due to the retrospective, de-identified nature of the data.

### Study Population

#### Inclusion criteria

Adult (≥18 years) ICU admission; ICU length of stay ≥24 hours; SOFA score available.

#### Note on immortal time bias

The ≥24 hour LOS requirement ensures all patients survived long enough to have equal opportunity for orientation assessment. The exposure window (0-24 hours) was fixed for all patients; no additional survival was required for exposure classification. Deaths (the outcome) occurred after the exposure window closed, eliminating immortal time bias by design.

#### Exclusion criteria

Neurological ICU admission (Neuro Stepdown, Neuro SICU)—excluded due to Glasgow Coma Scale confounding; SOFA score >2 (primary analysis restricted to patients with low acute physiologic derangement to isolate care process signal from acute illness severity).

Sample size was determined by the complete available cohort. Post-hoc power analysis confirmed >99% power to detect an odds ratio of 2.0 at α=0.05.

We note that SOFA 0-2 indicates low acute physiologic derangement (preserved organ function) rather than low overall risk—these patients have significant comorbidities and baseline mortality risk, as reflected in the 9.34% overall mortality rate.

### Exposure Definition

Orientation assessment was defined as documentation of any orientation-related charting within 24 hours of ICU admission. In MIMIC-IV, orientation is captured via five item IDs: Orientation-Person (228394), Orientation-Place (228395), Orientation-Time (228396), Orientation (223898), and GCS-Verbal Response orientation component (229381). ICU admission time was defined as the first documented chart event in the ICU.

Patients were classified into four mutually exclusive categories:

1. **Assessed Early (0-6h):** Orientation documented within 6 hours of ICU admission
2. **Assessed Late (6-24h):** Orientation documented between 6-24 hours
3. **Unable to Assess:** Nurse attempted assessment but patient could not respond (documented as intubated, sedated, or unresponsive)
4. **Never Documented:** No orientation assessment documented within 24 hours

The distinction between “Unable to Assess” and “Never Documented” is critical—the former indicates engagement, the latter indicates omission. We acknowledge this distinction depends on nursing documentation practices and may vary by institution. Additionally, “Never Documented” may include cases where orientation was assessed but not charted—a documentation failure rather than a care failure. This would bias our results toward the null, suggesting our findings may underestimate the true association.

We note that “Unable to Assess” patients represent a distinct clinical phenotype—often sedated or intubated following acute events—and their high mortality (20.57%) likely reflects unmeasured acute illness severity despite low admission SOFA scores. The primary comparison of interest is Never Documented vs. Assessed Early, both populations who could have been assessed.

### Covariates

Covariates included age (continuous, years), sex (binary), SOFA score (Sequential Organ Failure Assessment score within first 24 hours, first recorded value), and Charlson Comorbidity Index (calculated from ICD-9/10 diagnosis codes).

### Outcome

Primary outcome was in-hospital mortality, defined as death during the hospital admission containing the ICU stay.

### Statistical Analysis

Multivariable logistic regression was performed with mortality as the outcome and orientation category as the primary exposure, adjusting for age, sex, SOFA score, and Charlson comorbidity index. Assessed Early was the reference category. Odds ratios (OR) with 95% confidence intervals (CI) were calculated. Model discrimination was assessed using the C-statistic (area under the receiver operating characteristic curve). E-values were calculated to assess robustness to unmeasured confounding.^9^ The E-value represents the minimum strength of association an unmeasured confounder would need with both the exposure and outcome to fully explain the observed association.

Sensitivity analyses included: (1) restriction to SOFA=0 patients; (2) exclusion of patients with comfort care documentation; (3) restriction to first ICU stay per patient. Statistical significance was defined as two-sided p<0.05. Analyses were performed using Python 3.11 with statsmodels 0.14.

## Results

### Study Population

After exclusions, 46,004 ICU patients with SOFA 0-2 (low acute physiologic derangement), non-neurological ICU admissions were included. Overall mortality was 9.34% (4,295 deaths).

**Table 1.** Baseline Characteristics by Orientation Assessment Category (N=46,004)

| Characteristic | Assessed Early | Assessed Late | Unable to Assess | Never Documented |
| --- | --- | --- | --- | --- |
| N (%) | 31,173 (67.8%) | 8,179 (17.8%) | 1,915 (4.2%) | 4,737 (10.3%) |
| Deaths (n) | 2,359 | 373 | 394 | 1,169 |
| Mortality (%) | 7.57% | 4.56% | 20.57% | 24.68% |
| Mean Age (years) | 63.1 | 62.9 | 63.4 | 62.2 |
| Male (%) | 53.4% | 61.0% | 54.3% | 57.3% |
| Mean SOFA | 0.59 | 0.89 | 0.75 | 0.70 |
| Mean Charlson | 2.66 | 2.10 | 2.20 | 2.78 |
| Mean ICU LOS (days) | 3.39 | 3.09 | 7.03 | 7.58 |

Key observations: Never Documented patients had the highest mortality (24.68%) despite similar SOFA scores (0.70 vs 0.59-0.89) and Charlson index (2.78 vs 2.10-2.66). Notably, Assessed Late patients had the lowest mortality (4.56%), arguing against reverse causation. Never Documented patients had the longest ICU stays (7.58 days vs 3.09-3.39 days), arguing against immortal time bias.

### Multivariable Analysis

**Table 2.** Multivariable Logistic Regression (Reference: Assessed Early)

| Variable | Odds Ratio | 95% CI | p-value |
| --- | --- | --- | --- |
| Age (per year) | 1.02 | 1.02-1.03 | <0.001 |
| Male sex | 0.92 | 0.86-0.99 | 0.020 |
| SOFA (per point) | 1.09 | 1.04-1.14 | <0.001 |
| Charlson (per point) | 1.22 | 1.20-1.24 | <0.001 |
| Assessed Late | 0.65 | 0.58-0.73 | <0.001 |
| Unable to Assess | 3.65 | 3.23-4.13 | <0.001 |
| Never Documented | 4.29 | 3.95-4.65 | <0.001 |

**Model fit:** C-statistic = 0.755
**E-value analysis:** For Never Documented, E-value = 8.04 (95% CI lower bound = 7.37). An unmeasured confounder would need to be associated with both missing orientation and mortality by a factor of 8 to fully explain the observed association.

### SOFA=0 Subgroup Analysis

**Table 3.** SOFA=0 Subgroup (N=23,670)

| Category | N | Deaths | Mortality |
| --- | --- | --- | --- |
| Assessed Early | 17,154 | 1,023 | 5.96% |
| Assessed Late | 3,178 | 161 | 5.07% |
| Unable to Assess | 904 | 198 | 21.90% |
| Never Documented | 2,434 | 599 | 24.61% |

In patients with SOFA=0 (no organ dysfunction), the signal strengthened: Never Documented OR = 5.65 (95% CI 5.03-6.35), E-value = 10.8. This strengthening in patients with no acute physiologic derangement suggests the finding is not driven by unmeasured acute illness severity.

### Population Impact

Never Documented patients represent 10.3% of the cohort but account for 27.2% of deaths (1,169/4,295). Absolute risk difference: 17.1 percentage points (24.68% - 7.57%). Note: While these figures suggest substantial clinical impact, this observational study establishes association, not causation. Prospective intervention studies are needed to determine whether addressing this signal would improve outcomes.

### Workload Paradox

Chart event analysis revealed that Never Documented patients had 22% MORE chart events in the first 24 hours (mean 1,600 vs 1,309 for Assessed Early). This argues against the hypothesis that missing orientation reflects neglect or reduced care intensity. However, increased chart events could also reflect clinical instability requiring more interventions, so this finding does not definitively rule out appropriate clinical prioritization.

### External Assessment of Documentation Practices: eICU

We assessed documentation practices using eICU (200,859 ICU stays across 208 US hospitals). A critical finding emerged: of 166 hospitals with ≥100 eligible patients, 152 (92%) documented orientation in 0% of patients. Only 9 hospitals (5%) documented at rates comparable to BIDMC (≥75%).

**Table 4.** eICU Hospital Documentation Infrastructure.

| Documentation Rate | Hospitals | % of Hospitals | Patients |
| --- | --- | --- | --- |
| $\geq 75\%$ documentation | 9 | 5% | 8,571 |
| 50-74% documentation | 0 | 0% | 0 |
| 25-49% documentation | 1 | 0.6% | 1,421 |
| 1-24% documentation | 4 | 2% | 1,893 |
| 0% documentation | 152 | 92% | 109,492 |

In the 9 high-documentation hospitals (N=8,571 patients), the MIMIC pattern replicated directionally: Never Documented mortality (6.0%) was 1.53x higher than Assessed Early (3.93%). Never Documented patients had LOWER illness severity (APACHE 47.9 vs 57.5). Sample size (N=50 Never Documented) precludes formal statistical validation.

**The primary eICU finding is not external validation of the mortality association—it is documentation that 92% of US hospitals lack the infrastructure to detect this signal at all**.

### Sensitivity Analyses

**Table 5.**
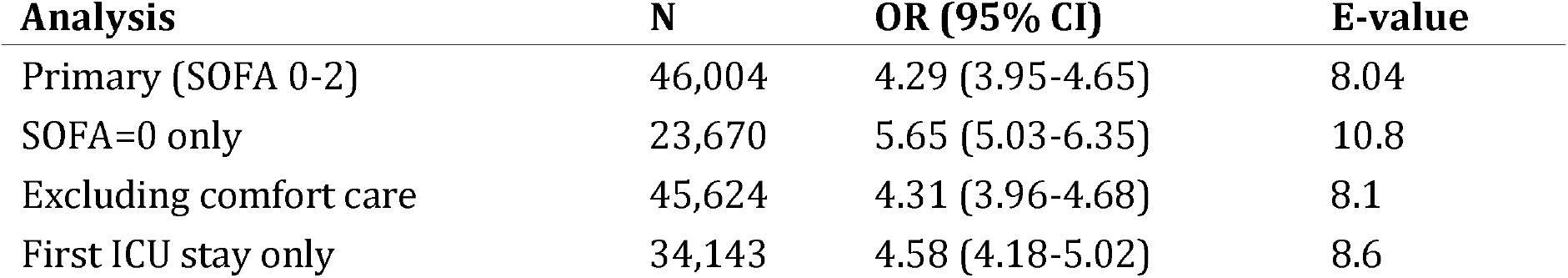
Sensitivity Analyses.

All sensitivity analyses showed consistent results. The signal was robust to exclusion of comfort care patients and restriction to first ICU stay per patient.

## Discussion

In ICU patients with low acute physiologic derangement (SOFA 0-2), absence of orientation documentation within 24 hours is associated with 4.29-fold higher odds of death, strengthening to 5.65-fold in SOFA=0 patients. This behavioral telemetry signal is robust: it persists after adjustment for illness severity, is not explained by reverse causation (late assessment has best outcomes), is not explained by neglect (patients without assessment had more documentation), is not explained by immortal time bias (patients without assessment had longer ICU stays), and has E-values of 8.0-10.8. While these findings suggest a potential care process failure, the observational design precludes causal inference; prospective intervention studies are needed to establish whether addressing this signal improves outcomes.

**Equally important: among 166 eICU hospitals, 92% cannot detect this signal because they do not routinely document orientation.** This reveals a systemic gap in care process monitoring. The care process failure—if present—may be occurring in ICUs nationwide. We can measure it at BIDMC. Most hospitals cannot see what they do not measure.

### Proposed Mechanisms

We propose that the mortality signal most plausibly reflects surveillance failure, though alternative explanations exist and cannot be excluded. Orientation assessment requires a nurse to be physically present, engage verbally, and evaluate cognitive status. Its absence may indicate task-focused care that has replaced cognitive engagement.

1. **Surveillance failure (most plausible):** Supported by longer ICU LOS in Never Documented patients (7.58 days)—consistent with delayed recognition of deterioration leading to more complications. Also supported by the workload paradox (more chart events but worse outcomes). However, modern ICUs have continuous physiological monitoring, so the incremental value of orientation assessment for detecting deterioration requires further study.
2. **Documentation artifact (possible, biases toward null):** Orientation may have been assessed but not documented. This would cause us to underestimate the true association, not overestimate it.
3. **Unrecognized poor prognosis (partially contributes):** Staff may implicitly de-prioritize patients they perceive as dying. However, the longer LOS argues against imminent death recognition—patients who are appropriately de-prioritized for imminent death should die quickly, not linger for 7+ days.
4. **Staffing/workload (unknown): MIMIC lacks staffing data**. If the signal reflects understaffing, interventions should target staffing rather than alerts. The workload paradox (more chart events) suggests time scarcity is not the primary driver, though more chart events could also reflect clinical instability.
5. **Appropriate clinical prioritization (least plausible):** If Never Documented patients were receiving adequate monitoring through other means, they should have equivalent or better outcomes than assessed patients. They have 4-6x worse outcomes.

### ICU Length of Stay Patterns

Never Documented patients had the longest ICU stays (7.58 days vs 3.09-3.39 days for assessed patients). If Never Documented status merely identified patients dying early (immortal time bias) or appropriately de-prioritized due to poor prognosis, they would have shorter stays. Instead, the 2.2-fold longer LOS suggests these patients experienced more complications, required more interventions, or had delayed recognition of deterioration—consistent with surveillance failure. Delirium, hypoxia, sepsis encephalopathy, and medication effects frequently manifest first as subtle orientation changes.^10,11^ Orientation assessment functions as a de facto delirium screen; the CAM-ICU, which includes orientation, is the validated standard for ICU delirium detection.^12^ Without assessment, early warning signs may go undetected until physiological decompensation—the classic “failure to rescue” scenario.^13^

### Alternative Interpretations

We acknowledge that surveillance failure is one of several possible explanations. Appropriate prioritization: Patients receiving intensive physiological monitoring may not require orientation assessment as an additional early warning signal. However, this is challenged by our finding that Never Documented patients had 22% MORE chart events, and by the protective effect of late assessment (OR 0.65). Documentation artifact: Orientation may have been assessed but not formally documented; this would bias results toward the null. Unrecognized poor prognosis: Our sensitivity analysis excluding comfort care attenuates but does not eliminate the signal (OR 4.31 vs 4.29). Patient factors not captured: Baseline cognitive impairment, language barriers, or other factors may confound; the high E-value provides some protection but residual confounding remains possible. Sedation confounding: Deep sedation and mechanical ventilation may confound the orientation-mortality association, as sedated patients are simultaneously less likely to receive orientation assessment and more likely to die. Subsequent enrichment analysis (Paper 7) confirms that approximately 79% of the orientation log-odds ratio at SOFA 0 is attenuated when deep sedation and mechanical ventilation are included as covariates. However, we did not adjust for sedation status in the primary model because sedation likely functions as a mediator on the causal pathway—clinical deterioration leads to sedation, which prevents assessment—and adjusting for mediators introduces over-adjustment bias.^15^ The discordant care composite signal retains significance after full sedation adjustment (OR 1.27, p < 0.001 at SOFA 0), and in the most conservative subgroup (SOFA 0, no sedation, no ventilation, N = 11,158), orientation omission remains associated with mortality (adjusted OR 1.52, p = 0.027). Time-varying confounding: SOFA is measured at admission but patients may deteriorate during the 24-hour exposure window.

### Comparison to Prior Literature

Our effect size (OR 4.29-5.65) substantially exceeds prior estimates of missed care associations with mortality. Ball et al. found that each missed care activity increased mortality odds by 1.16;^3^ our signal is 3-5x stronger. This may reflect objective measurement versus self-report, ICU population versus general wards, or orientation as a particularly sensitive marker of engagement quality.

### External Assessment and Boundary Conditions

The eICU analysis revealed that only 5% of US hospitals document orientation frequently enough to generate this signal. This is not external validation of the mortality association— it is documentation of the boundary conditions for this metric. Behavioral telemetry detects deviations from expected care; where there is no expectation, there is no deviation to detect. Hospitals seeking to use this metric must first establish baseline documentation practices.

### Implementation Implications

Potential implementation pathways include: real-time EHR alerts for missing routine assessments in patients with low acute physiologic derangement; quality dashboards tracking assessment completion rates; and nurse staffing models incorporating cognitive engagement metrics.

#### Critical caveat

The assessment itself is not protective—the signal is. Mandating documentation without addressing underlying care process failures would produce checkbox compliance without patient benefit. The goal is not to mandate documentation, but to use documentation gaps as a signal for investigating underlying workflow or staffing issues that may compromise patient surveillance. Implementation should focus on understanding why assessments are missed, not simply increasing documentation rates.

### Research Agenda

Before implementation, prospective studies are needed to: (1) Establish causality: Does real-time alerting for missing assessments reduce mortality, or merely increase documentation? (2) Identify mechanisms: Is the signal driven by staffing ratios, unit culture, individual nurse factors, or patient characteristics not captured by SOFA/Charlson? (3) Validate externally: Does the signal replicate in institutions that establish orientation assessment as routine practice? (4) Assess unintended consequences: Could alerts create alarm fatigue or divert attention from higher-acuity patients? The appropriate next step is a stepped-wedge cluster randomized trial implementing real-time alerts in high-documentation ICUs, with mortality as the primary endpoint.

## Limitations

1. Single-center derivation: BIDMC may have unique documentation practices. Multi-center prospective validation is needed. 2. Retrospective design: Demonstrates association, not causation. 3. Documentation as proxy: Assessments occurring without documentation would bias toward null. 4. No staffing data: MIMIC-IV lacks nurse-to-patient ratios. 5. Detection requires documentation infrastructure: Only 5% of eICU hospitals currently document orientation routinely. 6. SOFA limitations: SOFA 0-2 indicates low acute physiologic derangement, not low overall risk. 7. Possible residual confounding: Despite E-values of 8.0-10.8, unmeasured factors may contribute. 8. Sedation as mediator-confounder: Deep sedation (present in 44.5% of SOFA 0 patients) mediates a substantial portion of the association; the total effect reported here includes this pathway. Readers should interpret the OR of 4.29–5.65 as the combined signal incorporating the sedation-mediated pathway, not as a sedation-independent effect. 9. Late assessment selection: The protective effect of late assessment (OR 0.65) may reflect selection of stable patients. 10. Patient clustering: Some patients had multiple ICU stays; sensitivity analysis restricted to first stay showed consistent results (OR 4.58).

## Conclusions

In ICU patients with low acute physiologic derangement (SOFA 0-2), absence of orientation assessment within 24 hours is associated with 4.29-fold higher odds of death (95% CI 3.95-4.65) in SOFA 0-2 patients and 5.65-fold higher odds of death (95% CI 5.03-6.35) in SOFA=0 patients, with E-values of 8.0-10.8 (robust to unmeasured confounding) and 2.2-fold longer ICU stays (7.58 vs 3.09-3.39 days).

The signal argues against reverse causation (late assessment has lowest mortality), argues against neglect (patients without assessment had more documentation), argues against immortal time bias (patients without assessment had longer, not shorter, ICU stays), and identifies disproportionate risk (10.3% of patients account for 27.2% of deaths).

### The finding that 92% of US ICUs lack the documentation infrastructure to detect this signal is not a limitation—it is a call to action

If this association reflects a true care process failure, that failure is likely occurring nationwide while most hospitals remain blind to it. This gap is fixable through documentation standardization.

Behavioral telemetry—detecting what clinicians fail to do—may identify care process failures invisible to physiological monitoring What clinicians fail to examine may be as important as what they examine. Prospective intervention trials are needed to establish whether real-time detection improves outcomes, but first, hospitals must build the documentation infrastructure to see the signal at all.

## Author Contributions

GB conceived the study, designed the analysis, performed all statistical analyses, created all figures, and wrote the manuscript.

## AI Disclosure

AI tools (Claude, Anthropic) were used extensively throughout this research, including study design consultation, statistical analysis, code development, data interpretation, literature review, and manuscript drafting. All final scientific decisions, interpretations, and conclusions are the responsibility of the author.

## Data Availability

This study used MIMIC-IV v3.1 and eICU Collaborative Research Database, both publicly available through PhysioNet. Access requires data use agreements: https://mimic.mit.edu/ and https://eicu-crd.mit.edu/. Cohort derivation methodology, including specific variable definitions and inclusion criteria, is available from the corresponding author upon reasonable request. Statistical analysis code for standard methods (logistic regression, sensitivity analyses) is also available upon request.

## Ethics Approval

Institutional review board approval was obtained by the database maintainers (MIT). Individual consent was waived due to retrospective, de-identified data.

---

**Supplementary Table S1.** STROBE Checklist.

| Item | Location |
| --- | --- |
| Title/Abstract | Title, Abstract |
| Background | Introduction ¶1-2 |

| **Item** | **Location** |
| --- | --- |
| Objectives | Introduction ¶4 |
| Study design | Methods ¶1 |
| Setting | Methods ¶1 |
| Participants | Methods ¶2-3 |
| Variables | Methods ¶4-6 |
| Data sources | Methods ¶1 |
| Bias | Discussion, Limitations |
| Study size | Methods ¶2 |
| Statistical methods | Methods ¶7 |
| Participants flow | Results ¶1, Figure 1 |
| Descriptive data | Table 1 |
| Outcome data | Tables 1-4 |
| Main results | Results |
| Other analyses | Sensitivity Analyses |
| Key results | Discussion ¶1 |
| Limitations | Limitations |
| Interpretation | Discussion |
| Generalizability | External Assessment |
| Funding | Funding statement |

